# Genetic predisposition to raised circulating levels of dietary antioxidants and the association with respiratory health in high-risk populations

**DOI:** 10.1101/2024.12.13.24318986

**Authors:** A Saied, L. J. Horsfall

**Affiliations:** Research Department of Primary Care and Population Health, University College London, UK; Institute of Health Informatics, University College London, UK

**Keywords:** antioxidant, diet, lung function, Mendelian randomization

## Abstract

**Background:** Observational studies of raised dietary antioxidants suggest a beneficial effect on health, but the results from interventional studies generally show no effect. There are no robust studies targeting people exposed to high levels of environmental oxidants where any effects of raised antioxidants are plausibly stronger.

**Objectives:** To examine whether people genetically predisposed to raised dietary serum antioxidants have improved respiratory health.

**Methods:** We identified single-nucleotide polymorphisms (SNPs) associated with serum ascorbate (vitamin C), retinol (vitamin A), and β-carotene from external data sources. Outcome data on forced expiratory volume in one second (FEV_1_) and forced vital capacity (FVC), were derived from individual-level data from over 285,000 UK Biobank participants. We used linear regression to explore relationships between the SNPs and lung function. To quantify any relationships, we instrumented the association using exposure beta coefficient from published literature and genome wide association studies (Mendelian randomization).

**Results:** We found no consistent relationship between genetically raised serum antioxidant levels and respiratory health measures. There was no evidence of effect modification by exposures linked to oxidative stress including cigarette smoke, air pollution and poor diet.

**Conclusions:** Our findings support interventional studies showing no causal relationship between dietary antioxidants and respiratory disease outcomes. Further, our results do not support interventions to increase serum levels of ascorbate, retinol, or β-carotene in people exposed to high levels of environmental oxidants (Wellcome Grant ID: 209207/Z/17/Z and 225195/Z/22/Z).

## Introduction

Chronic Obstructive Pulmonary Disease (COPD) is the fourth most common cause of death in the world and it contributes significantly to the worldwide burden of disease with an estimated global prevalence of 10.6%^1^. Although smoking tobacco is known to be the first causal and most important risk factor for COPD, there is a growing body of evidence which links diet with an accelerated decline of pulmonary function and an increased risk of COPD^2^. Spirometry is a key component of lung function tests used to diagnose and monitor respiratory conditions like COPD.

Oxidative stress is an imbalance between oxidants (reactive oxygen and nitrogen species) and antioxidants that causes damage to tissues, proteins, and DNA. Reactive oxygen and nitrogen species within the lung are mainly produced by phagocytes and polymorphonuclear, alveolar, bronchial, and endothelial cells. The lungs are also continuously exposed to exogenous oxidants such as cigarette smoke, mineral dust, ozone, and radiation. Oxidative stress has been shown to have a role in the pathogenesis of diffuse lung diseases and it has been hypothesized that increases in antioxidants to scavenge free radicals will protect against lung disease^3 4^.

Supporting this hypothesis, observational studies have found a link between antioxidant intake and improved lung function^3 5-9^. Serum levels of vitamins A, C, D, and α-tocopherol vitamin E are also associated with increased respiratory morbidity and/or mortality^10^. However, it is recognized that the ability to infer causal inference from this study design is limited due to biases such as residual confounding and reverse causation^11^.

Randomized controlled trials (RCTs) would be more helpful in verifying causal relationships between antioxidant use and lung function. Overall systematic reviews of RCTs have failed to identify protective effects of antioxidant vitamins against cancer, cardiovascular disease and death seen in observational studies, with some reporting an elevated risk of certain disease (e.g. β-carotene and lung cancer)^12^. For COPD prognosis, the evidence is less clear with a recent systematic review of RCTs finding high-dose vitamin C supplementation has a beneficial effect on lung function^13^. There are, however, important limitations of RCTs of dietary antioxidants, including small sample sizes, intervention doses that greatly exceed the recommended daily allowance, and the short duration of exposure. Furthermore, it is biologically plausible that supplementation may only benefit people with inadequate antioxidant levels but most RCTs do not target these high-risk subgroups. For example, people who smoke cigarettes are exposed to high levels of oxidative stress and may benefit more physiologically from supplementation than non-smokers. An RCT of the antioxidant selenium tested this hypothesis and found a stronger protective effect against lung function decline in smokers^14^ . There are few other identifiable studies that have tested this hypothesis *a priori*. Given the limitations of the observational and interventional evidence base, other study designs would be valuable for clarifying the role of long-term antioxidant modification in lung disease etiology.

Here, we present a large-scale study examining the effect of genetically raised levels of ascorbic acid (vitamin C), retinoic acid (vitamin A), and β-carotene on measures of lung function in adults participating in the UK Biobank. Using genetic polymorphisms to instrument serum concentrations of antioxidants, via a Mendelian randomization (MR) approach, can provide robust causal evidence provided strong assumptions are met. We hypothesized that any benefit of these variants on lung function would be particularly apparent for people exposed to higher levels of oxidative stress through smoking, diet or air pollution.

## Materials and methods

### Data Source

UK Biobank is a cohort containing genotype data matched with deep phenotype data for 500,000 volunteer participants aged between 40 and 69, living in the UK^15^. Participants were recruited between April 2007 and August 2010 and provided data on their health using questionnaires, physical measurements by a trained nurse, and blood samples. Genome-wide genotype data were available for the Affymetrix UK Biobank Axiom® array (most participants) and the Applied Biosystems™ UK BiLEVE Axiom™ Array by Affymetrix (a smaller subset of n=49,950). Details on the quality control and imputation of SNPs, indels and structural variants are reported elsewhere^15^. Online tools were used to check the adequacy of the sample size (http://cnsgenomics.com/shiny/mRnd/).

### Study design

First, we investigated the cross-sectional relationships between SNPs associated with circulating antioxidant levels and lung function. We then instrumented the strength of the association using published summary statistics (beta coefficients) for serum antioxidants and individual level lung function data measured in UK Biobank participants at baseline (two-sample MR). The present study protocol was pre-approved by UK Biobank (ID: 5167). A shorter version of this work was presented as a poster at the European Society for Human Genetics and published as an abstract^16^. The present manuscript is longer, contains a detailed literature review, extensive methodology, additional results, in-depth analyses, and a thorough discussion of results^17^.

### Inclusion/exclusion criteria

The number of participants in UK Biobank is 502,527. We excluded outliers for genotype missingness, excess heterozygosity, and sex discordance (n=2,200). We also removed close relatives to ensure participants had no close family member in the study (n=39,642). We further restricted the sample to ‘white British’ participants using the results of an existing principal components analysis and participants self-reported ethnicity (n=88,341).

#### Genetic instruments

We considered genetic variants as instrumental variables for three circulating antioxidants: ascorbic acid (vitamin C), retinol (vitamin A), and β-carotene. We used the NHGRI-EBI GWAS catalog (http://www.ebi.ac.uk/gwas/) and a wider literature search using the PubMed search engine to identify unlinked SNPs most strongly associated with the circulating trait of interest. The SNP identified as an instrument for ascorbate is a missense variant in an exon of Solute Carrier Family 23 Member 1 (SLC23A1), which encodes sodium-dependent vitamin C transporter 1 (SVCT1), 1 of 2 cotransporters involved in the intestinal absorption and active transport of dietary ascorbate^18^ . The SNPs instrumenting β-carotene is located in beta-carotene oxygenase 1 (BCO1/alias BCMO1), which encodes for an enzyme which catalyzes the cleavage of carotenoids into retinal in the small intestine (carotenoid 15,15-monooxygenase)^19^ . The SNP instrumenting retinol is located near retinol-binding protein 4 (RBP4), responsible for retinol transport from liver stores to peripheral tissues. Deleterious mutations in RBP4 have been associated with biochemical vitamin A deficiency and related disorders, such as retinitis pigmentosa^20^.

#### Outcomes

The primary outcome is lung function as measured as forced expiratory volume in one second (FEV_1_) and the forced vital capacity (FVC). Breath spirometry was performed on participants attending the assessment center using a Vitalograph Pneumotrac 6800. The participant was asked to record two to three blows (lasting for at least six seconds) within a period of approximately 6 minutes. The computer then compared the reproducibility of the first two blows and, if acceptable (defined as a <5% difference in FVC and FEV_1_), indicated that the third blow is not required. Our previous work has found that introducing these types of quality control to lung function can exclude people with the lowest lung function and may introduce selection bias. Therefore, we also analyzed the best of any blow recorded without exclusions based on acceptability as a sensitivity analysis.

#### Covariates

To improve the precision of our estimates, known predictors of variation in lung function were included in our models. These were age, genetic sex, height, weight, and self-reported smoking status. We also included recruitment center and the first 40 principal components to help adjust for any population sub-structure within the “white British” participants. We tested for interactions with variables that might influence levels of oxidative stress including smoking status (categorical variable; never, former and current), pack-years of smoking (continuous variable), daily raw fruit and vegetable intake (continuous variable), daily dietary supplements containing purported antioxidants (binary), and area-linked levels of NO_2_ air pollution (continuous).

#### Statistical analyses

We identified and excluded outlier continuous values using a multivariate approach (blocked adaptive computationally efficient outlier nominators algorithm) with a 15% threshold of the chi-squared distribution used to separate outliers from non-outliers. We used linear regression to estimate the associations between genotypes and lung function after including important predictors (e.g., sex at birth) and potential confounders (e.g., principal components). To visualize potential gene-environment interactions, we calculated the margins of response as adjusted FEV_1_ and FVC values in liters for different levels of environmental exposures linked to oxidative stress (smoking, air pollution, poor diet) while holding all other variables at their observed values. We used the Wald test to calculate the p-values associated with interaction terms. To estimate the causal effect of a unit increase in each circulating antioxidant on lung function, we extracted beta coefficients of the SNP-exposure from the literature and combined these with the beta coefficients for SNP-outcome from UK Biobank using the Wald ratio^21^. We performed several sensitivity analyses to test the robustness of the findings. Lung function was missing for approximately 25% of participants, and missingness was associated with several other risk factors, such as smoking status, and therefore not at random. As a sensitivity analysis, we imputed missing lung function and other continuous variables (height, weight) using multivariate normal regression and recalculated the associations. This method uses an iterative Markov chain Monte Carlo method to impute missing values, and we used n⍰= ⍰10 imputations. Recent use of respiratory medication was self-reported at baseline. These medications could affect spirometry readings, so we also assessed the impact of excluding participants reporting to be on these drugs.

Statistical analyses were completed using Stata version 16.1 and R version 4.2.2.

## Results

Approximately 285,000 participants had data on both genetic variants of interest and lung function (Table 1). Mean FEV_1_ was 2.85 (SD ±0.77) and mean FVC was 3.78 (SD ±0.97). There was no clear association between alleles associated with increased retinol or β-carotene and increased lung function (Table 1). In participants with alleles associated with raised ascorbate, FEV_1_ was 9 ml higher and FVC was 10 ml higher. The unadjusted and adjusted coefficients show a positive association between alleles associated with increased circulating antioxidants and lung function for most analyses (Table 2). The positive association between alleles increasing ascorbate levels and lung function reversed after adjustment. For example, before adjustment, each additional effect allele was associated with 11.7 ml increase in FEV_1_ (95%CI: 1.6 to 21.9) which reduced to 1.3 ml (-6.0 to 8.6) on adjustment. Further analyses found that a slight imbalance in sex and height across ascorbate genotypes explained the positive relationship suggested by Table 1 and the unadjusted analyses in Table 2. Imputing missing data for continuous variables substantially increased our analytic sample but did not alter our findings (Table 2). We found no clear evidence of interactions between genotypes and smoking status in the regression models (Table 3) or the results of the two-sample MR (Figure 1). There was also no evidence of interactions with pack-years of smoking, air pollution levels, vitamin supplements or intake of raw fruit and vegetables.

**Table 1.** Characteristics of UK Biobank participants included in the analysis overall and by number of effect alleles.

| rs4889286 | Number of effect alleles |  |  |  | n | Missingness |
| --- | --- | --- | --- | --- | --- | --- |
|  | Total | 0 effect alleles | 1 effect alleles | 2 effect alleles |  |  |
| (β-carotene) | N=374,924 | N=82,766 | N=186,850 | N=105,308 | 374,924 | |
| Male sex | 173,644 (46.3%) | 38,237 (46.2%) | 86,560 (46.3%) | 48,847 (46.4%) | 374,924 | 0.00% |
| Age at recruitment* | 58.9 (51.4-64.0) | 58.9 (51.3-64.0) | 58.9 (51.4-63.9) | 58.9 (51.3-64.0) | 374,924 | 0.00% |
| Weight (kg) | 78.3 (15.5) | 78.3 (15.5) | 78.3 (15.4) | 78.2 (15.4) | 373,875 | 0.28% |
| Height (cm) | 168.8 (9.1) | 168.8 (9.2) | 168.8 (9.1) | 168.8 (9.1) | 374,166 | 0.20% |
| Smoking status |  |  |  |  | 374,924 | 0.00% |
| Never | 204,092 (54.4%) | 45,048 (54.4%) | 101,851 (54.5%) | 57,193 (54.3%) |  |  |
| Former | 131,731 (35.1%) | 29,141 (35.2%) | 65,502 (35.1%) | 37,088 (35.2%) |  |  |
| Current | 37,822 (10.1%) | 8,314 (10.0%) | 18,849 (10.1%) | 10,659 (10.1%) |  |  |
| Missing | 1,279 (0.3%) | 263 (0.3%) | 648 (0.3%) | 368 (0.3%) |  |  |
| FEV<sub>1</sub> | 2.85 (0.77) | 2.85 (0.77) | 2.85 (0.76) | 2.85 (0.77) | 284,486 | 24.12% |
| FVC | 3.78 (0.97) | 3.78 (0.97) | 3.78 (0.97) | 3.78 (0.97) | 284,486 | 24.12% |
| rs10882272 | Total | 0 effect alleles | 1 effect alleles | 2 effect alleles | n | Missingness |
| (Retinol) | N=376,771 | N=54,228 | N=176,918 | N=145,625 | 376,771 |  |
| Male sex | 174,538 (46.3%) | 24,937 (46.0%) | 82,392 (46.6%) | 67,209 (46.2%) | 376,771 | 0.00% |
| Age at recruitment | 58.9 (51.3-64.0) | 58.9 (51.3-64.0) | 58.9 (51.4-63.9) | 58.9 (51.3-64.0) | 376,771 | 0.00% |
| Weight (kg) | 78.3 (15.5) | 78.1 (15.4) | 78.3 (15.5) | 78.3 (15.5) | 375,715 | 0.28% |
| Height (cm) | 168.8 (9.1) | 168.7 (9.1) | 168.9 (9.1) | 168.8 (9.2) | 376,010 | 0.20% |
| Smoking status |  |  |  |  | 376,771 | 0.00% |
| Never | 205,088 (54.4%) | 29,460 (54.3%) | 96,403 (54.5%) | 79,225 (54.4%) |  |  |
| Former | 132,373 (35.1%) | 19,030 (35.1%) | 62,073 (35.1%) | 51,270 (35.2%) |  |  |
| Current | 38,022 (10.1%) | 5,562 (10.3%) | 17,842 (10.1%) | 14,618 (10.0%) |  |  |
| Missing | 1,288 (0.3%) | 176 (0.3%) | 600 (0.3%) | 512 (0.4%) |  |  |
| FEV<sub>1</sub> | 2.85 (0.77) | 2.85 (0.77) | 2.86 (0.77) | 2.85 (0.76) | 285,877 | 24.12% |
| FVC | 3.78 (0.97) | 3.77 (0.97) | 3.79 (0.97) | 3.78 (0.97) | 285,877 | 24.12% |
| rs33972313 | Total | 0 effect alleles | 1 effect alleles | 2 effect alleles | n | Missingness |
| (Ascorbate) | N=376,771 | N=459 | N=25,454 | N=350,858 | 376,771 |  |
| Male sex | 174,538 (46.3%) | 188 (41.0%) | 11,716 (46.0%) | 162,634 (46.4%) | 376,771 | 0.00% |
| Age at recruitment | 58.9 (51.3-64.0) | 57.6 (49.8-62.8) | 58.9 (51.2-64.0) | 58.9 (51.4-64.0) | 376,771 | 0.00% |
| Weight (kg) | 78.3 (15.5) | 77.8 (15.5) | 78.2 (15.5) | 78.3 (15.4) | 375,715 | 0.28% |
| Height (cm) | 168.8 (9.1) | 167.9 (8.7) | 168.7 (9.1) | 168.8 (9.2) | 376,010 | 0.20% |
| Smoking status |  |  |  |  | 376,771 | 0.00% |
| Never | 205,088 (54.4%) | 239 (52.1%) | 13,827 (54.3%) | 191,022 (54.4%) |  |  |
| Former | 132,373 (35.1%) | 166 (36.2%) | 8,989 (35.3%) | 123,218 (35.1%) |  |  |
| Current | 38,022 (10.1%) | 51 (11.1%) | 2,562 (10.1%) | 35,409 (10.1%) |  |  |
| Missing | 1,288 (0.3%) | 3 (0.7%) | 76 (0.3%) | 1,209 (0.3%) |  |  |
| FEV<sub>1</sub> | 2.85 (0.77) | 2.76 (0.76) | 2.85 (0.76) | 2.85 (0.77) | 285,877 | 24.12% |
| FVC | 3.78 (0.97) | 3.68 (0.94) | 3.78 (0.96) | 3.78 (0.97) | 285,877 | 24.12% |

\*Median with interquartile range

**Table 2.**
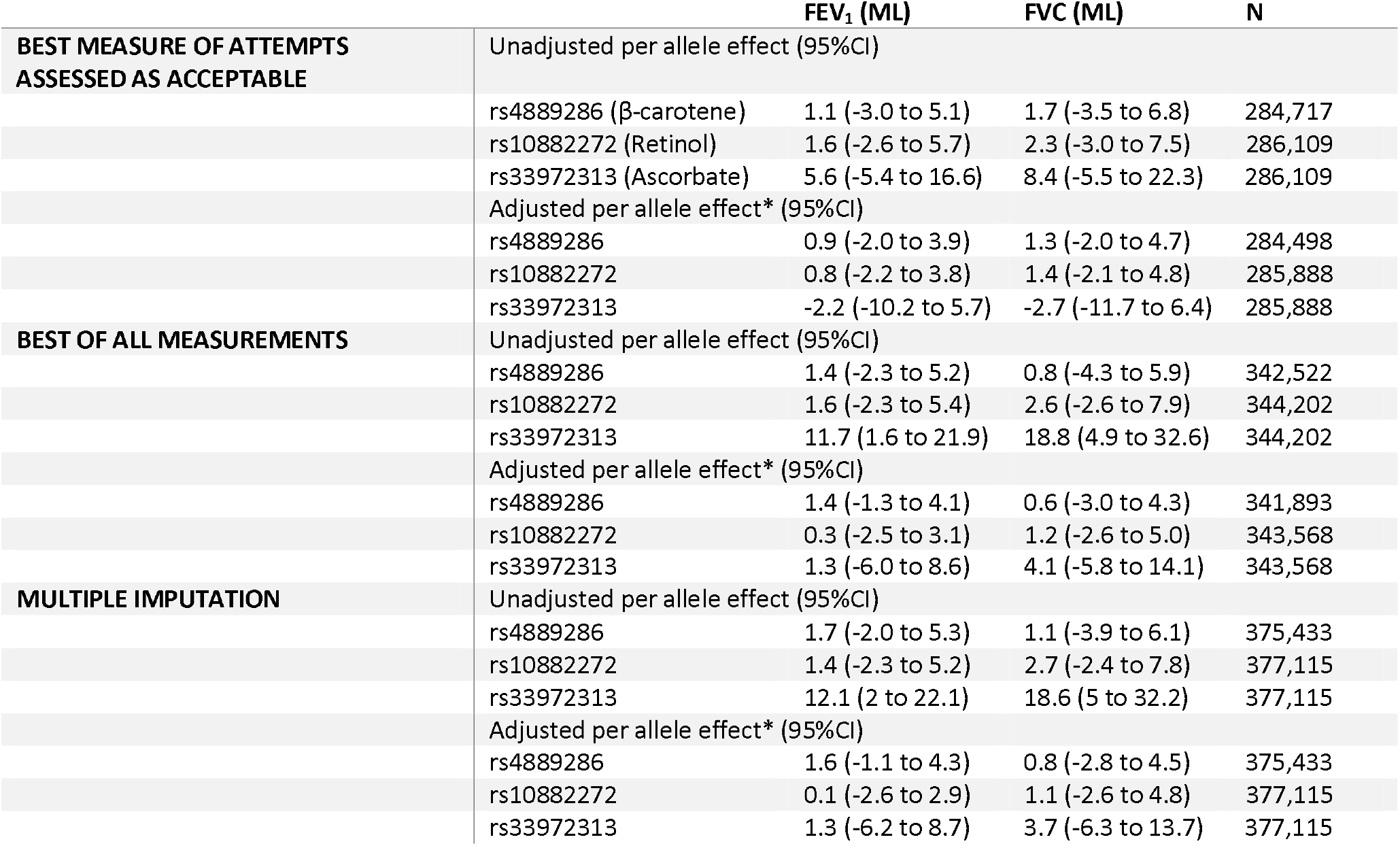
Genetic variants influencing circulating dietary antioxidants and their association with lung function in UK Biobank participants.

**Table 3.** Interaction between smoking status and Genetic variants influencing circulating dietary antioxidants and their association with lung function.

| *Smoking status* | *No. effect alleles* | *rs4889286*<br>*(β-carotene)* | *rs10882272*<br>*(Retinol)* | *rs33972313*<br>*(Ascorbate)* |
| --- | --- | --- | --- | --- |
| Predicted FEV<sub>1</sub> in litres (95% CI)* |  |  |  |  |
| *Never* | 0 | 3.83 (3.82 to 3.83) | 3.82 (3.82 to 3.83) | 3.77 (3.68 to 3.86) |
|  | 1 | 3.83 (3.82 to 3.83) | 3.83 (3.82 to 3.83) | 3.83 (3.82 to 3.84) |
|  | 2 | 3.83 (3.82 to 3.83) | 3.83 (3.82 to 3.83) | 3.83 (3.82 to 3.83) |
| *Former* | 0 | 3.74 (3.73 to 3.74) | 3.74 (3.73 to 3.75) | 3.74 (3.63 to 3.84) |
|  | 1 | 3.74 (3.74 to 3.75) | 3.74 (3.74 to 3.75) | 3.74 (3.73 to 3.76) |
|  | 2 | 3.74 (3.73 to 3.75) | 3.74 (3.73 to 3.74) | 3.74 (3.74 to 3.74) |
| *Current* | 0 | 3.70 (3.69 to 3.72) | 3.71 (3.69 to 3.72) | 3.91 (3.71 to 4.11) |
|  | 1 | 3.71 (3.70 to 3.72) | 3.71 (3.70 to 3.72) | 3.72 (3.69 to 3.75) |
|  | 2 | 3.72 (3.71 to 3.73) | 3.72 (3.70 to 3.73) | 3.71 (3.70 to 3.72) |
| *P for interaction* |  | 0.50 | 0.27 | 0.12 |
| Predicted FVC in litres (95% CI)* |  |  |  |  |
| *Never* | 0 | 2.92 (2.91 to 2.92) | 2.91 (2.91 to 2.92) | 2.87 (2.79 to 2.95) |
|  | 1 | 2.92 (2.91 to 2.92) | 2.92 (2.91 to 2.92) | 2.92 (2.91 to 2.93) |
|  | 2 | 2.92 (2.91 to 2.92) | 2.92 (2.91 to 2.92) | 2.92 (2.91 to 2.92) |
| *Former* | 0 | 2.80 (2.79 to 2.8) | 2.80 (2.79 to 2.81) | 2.77 (2.67 to 2.86) |
|  | 1 | 2.80 (2.80 to 2.81) | 2.80 (2.80 to 2.81) | 2.80 (2.79 to 2.82) |
|  | 2 | 2.80 (2.80 to 2.81) | 2.80 (2.79 to 2.81) | 2.80 (2.80 to 2.80) |
| *Current* | 0 | 2.69 (2.68 to 2.71) | 2.70 (2.68 to 2.72) | 2.83 (2.65 to 3.02) |
|  | 1 | 2.70 (2.69 to 2.71) | 2.70 (2.69 to 2.71) | 2.71 (2.68 to 2.73) |
|  | 2 | 2.71 (2.7 to 2.72) | 2.71 (2.70 to 2.72) | 2.70 (2.69 to 2.71) |
| *P for interaction* |  | 0.29 | 0.24 | 0.34 |

\*Adjusted for sex, height, weight, ethnicity principal components, smoking, and recruitment center.

**Figure 1.**
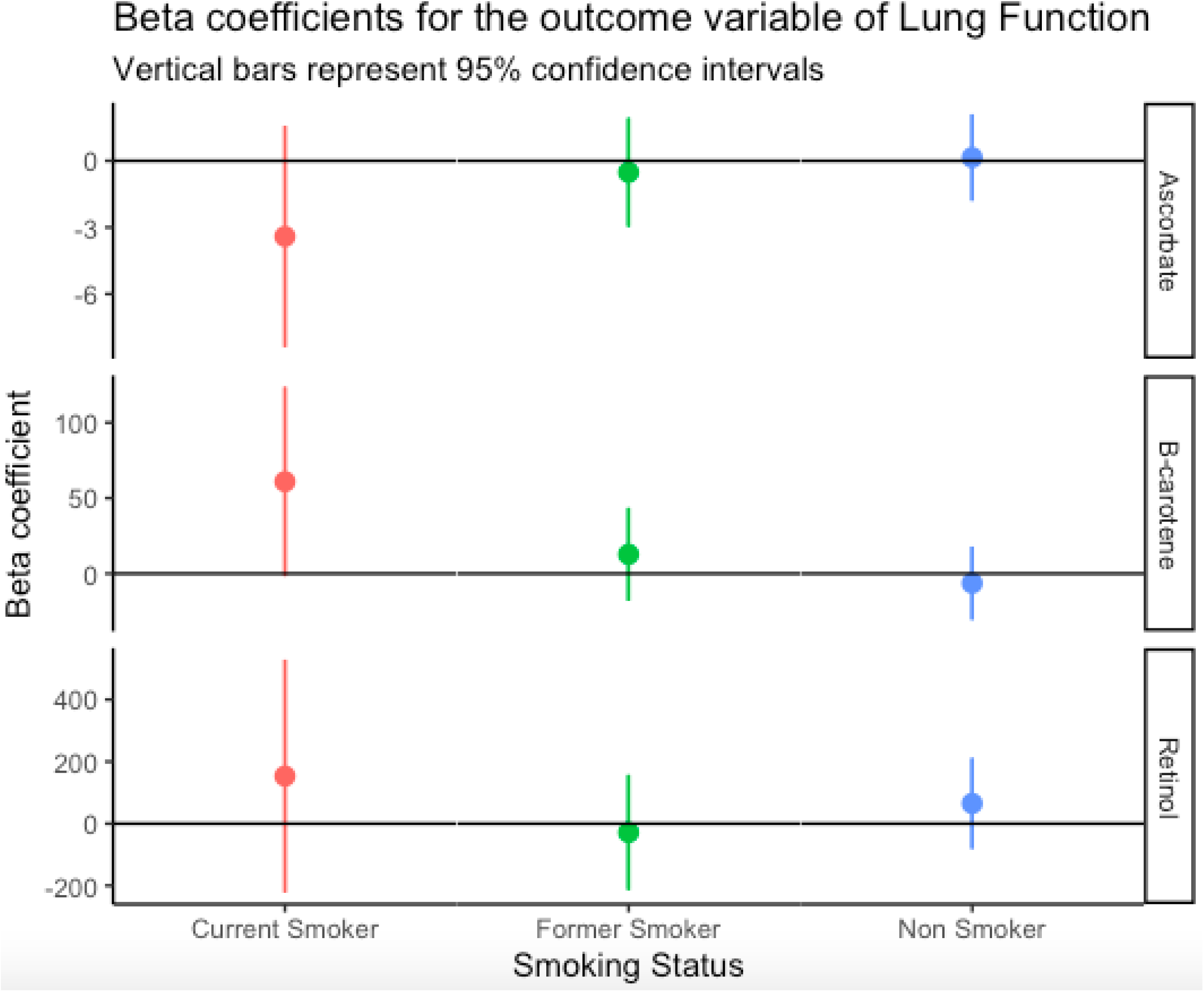
MR results for antioxidant exposure and lung function according to self-reported smoking status. β coefficients and 95% confidence intervals show unit difference in FVC per μmol/L for ascorbate, and unit difference FVC per natural log μg/L of serum β-carotene and serum retinol.

## Discussion

### Summary

This study examined whether genetically raised circulating dietary antioxidants are associated with improved lung function. Although most relationships were positive, there was limited evidence to support a clear association overall or in subgroups exposed to cigarette smoke, nitrogen dioxide air pollution or diets low in antioxidants. This suggests that long-term exposure to slightly raised serum ascorbate (vitamin C), retinol (vitamin A), and β-carotene are unlikely to protect lung health in a clinically meaningful way even in people exposed to high levels of oxidants.

### Strengths and Limitations

The main strength of this study was the use of genetic instruments to reduce bias from reverse causation and residual confounding that can be present in studies using measured blood levels of antioxidants. Other strengths include the large sample size and the ability to look at subsets of participants exposed to higher levels of oxidants, which is not usually possible with MR analyses using summary-level data.

Limitations include self-reporting of smoking status and other variables used to estimate exposure to oxidative stress. Exposure misclassification and poor measurement may have reduced our ability to detect important interactions. We lacked statistical power to examine interactions with clinical endpoints such as lung cancer diagnosis or mortality. It is recognized that there is selection bias among people who volunteer to join the UK Biobank, with smokers and people with known respiratory diseases significantly underrepresented^22 23^. A true causal association might be diluted if people with genetically low antioxidants are less likely to participate in UK Biobank due to death or poor health^24^. Furthermore, because the GWAS studies were based on white Europeans, we only restricted our analysis to white British participants. This limits the generalizability of our findings. When the instruments were selected for this study, we identified just one unlinked biologically plausible SNP for each antioxidant of interest. This restricts the power of our analyses and inhibits our ability to perform additional checks for evidence of bias due to horizontal pleiotropy. As more GWAS studies become available on circulating dietary antioxidants, the instruments can be expanded and precision to estimate causal effects increased.

There may also be pleiotropic effects that violate the third condition of an instrumental variable. The instrument for raised serum vitamin C, the SNP at location rs33972313, was weakly associated with height. On average, participants with effect alleles were almost 1cm taller (167.9cm vs 168.8cm). Two otherwise identical participants at these heights would expect a 1.5% difference in their FVC measurements. It seems biologically plausible that someone with higher levels of dietary-derived nutrients in their blood might be taller, anatomically increasing lung function independently of an antioxidant mechanism. It is also plausible that our selected SNPs influence levels of other antioxidants or metabolites. For example, there is some evidence that the SNP that increases serum levels of B-carotene decreases serum levels of other carotenoids^19^.

### Comparison with other studies

Our findings are consistent with existing RCTs and do not strongly support the previous observational studies that found a link between antioxidants and health^3 5-10^. These observational studies are prone to systematic biases such as residual confounding. This occurs between people who take supplemental antioxidants, other health-protective behaviors, and higher socioeconomic status. Another bias is reverse causation, where people possibly change their antioxidant intake after becoming unwell. Additionally, lung damage might affect how the body utilizes circulating antioxidants.

RCTs have failed to replicate these findings. The largest and most relevant is the heart protection study conducted in 2002^25^, where 20,536 UK adults with coronary disease, other occlusive arterial diseases, or diabetes were randomly allocated to receive antioxidant vitamin supplementation (600 mg vitamin E, 250 mg vitamin C, and 20 mg β-carotene daily) or matching placebo. As a secondary outcome measure, respiratory function was assessed by spirometry in all those attending the final follow-up visit. There were no significant differences in forced expiratory volume during one second observed between the treatment groups (FEV_1_: 2·06 L vitamin-allocated vs 2·06 L placebo-allocated; difference 0·00 L [SE 0·01]) or in forced vital capacity (FVC: 2·83L vs 2·82L; difference 0·01 L [SE 0·01]). However, the intervention doses were much larger than people would reasonably be expected to consume daily. The heart protection study gave 250mg of vitamin C daily, compared to the recommended daily allowance (RDA) of 75-90mg/day.

Similar MR studies have been used to investigate the effects of circulating levels of antioxidants on risk of coronary heart disease^26^ Alzheimer’s disease^27^, ischemic stroke^28^, prostate cancer^29^ and six major mental disorders^30^. While not all MR studies of antioxidants produce null findings^31-34^, most high-quality analyses fail to find any evidence of protective association despite being well-powered.

### Future directions

Understanding the causal relationship between dietary antioxidants is a vital area for research given the low cost of intervention and the high burden of oxidative-stress mediated disease. Future large-scale MR studies with multiple genetic instruments and repeated lung function measures for participants recruited at a younger age to reduce selection bias will improve the causal evidence base on dietary antioxidants. Well-designed RCTs investigating the effect of recommended daily amounts of antioxidant vitamins on lung function in young people living in areas with high concentrations of airborne oxidants would also be a valuable addition to the evidence base.

## Conclusion

We found no clear relationship between genetic variants influencing serum dietary antioxidant levels and lung function overall or in people exposed to higher levels of oxidative stress. In conclusion, this study together with existing evidence-base cast doubt on dietary antioxidants as a modifiable factor for improved lung health even in high-risk populations.

## Author Contributions

Conceptualization, A.S. and L.J.H.; methodology, A.S. and L.J.H.; software, A.S. and L.J.H.; .formal analysis, L.J.H.; investigation A.S. and L.J.H.; resources, L.J.H.; data curation, L.J.H.; writing—original draft preparation, A.S.; writing—review and editing, L.J.H.; visualization, A.S.,: supervision, L.J.H..; project administration, L.J.H.; funding acquisition, L.J.H.; All authors have read and agreed to the published version of the manuscript.

## Ethics statement

UK Biobank has ongoing approval from the North West Multi-centre Research Ethics Committee as a Research Tissue Bank (RTB) approval. This approval means that researchers do not require separate ethical clearance and can carry out research under the RTB approval. The present study protocol has been approved by UK Biobank (ID: 5167).

## Data Availability Statement

The data supporting this study’s findings are available from UK Biobank (https://www.ukbiobank.ac.uk/enable-your-research/apply-for-access). Restrictions apply to the availability of these data under Material Transfer Agreements.

## Acknowledgments

This research uses data from UK Biobank, a major biomedical database: www.ukbiobank.ac.uk. We want to thank the participants for contributing to this valuable resource.

